# Dynamics, Optimal Control, and Spillover Risk of the 2026 Bundibugyo Ebola Outbreak in the Democratic Republic of the Congo

**DOI:** 10.64898/2026.08.17.26360567

**Authors:** Jiahui Li, Shengjie Lai, Yanhua Su, Quipping Chen, Jia Rui, Zeyu Zhao, Tianmu Chen

## Abstract

In 2026, a Bundibugyo ebolavirus (BDBV) outbreak emerged in the Democratic Republic of the Congo (DRC), with 4,566 confirmed cases and 2,128 deaths reported as of 11 August, potentially becoming the largest Ebola outbreak on record globally. We developed a susceptible-exposed-infectious-deceased-recovered (SEIDR) model incorporating incorporating three categories of interventions—public self-protection, safe burial, and treatment and convalescence—to assess early transmission dynamics, the current epidemic trajectory, and cross-border spillover risk, and to inform the formulation of control strategies. Based on cumulative confirmed case data up to 31 July, sensitivity analyses across multiple candidate start dates identified 28 March as the optimal start date of sustained transmission, with 31 March to 3 April as the most likely onset window. As of 31 July, the basic reproduction number (ℛ_0_) was 1.83 (95% CI: 1.81–1.84). When 58.12% of the susceptible population adopted protective behaviours, the transmission chain could be effectively interrupted. By integrating the non-dominated sorting genetic algorithm II (NSGA-II) with Pontryagin’s minimum principle (PMP), we derived a time-varying optimal control strategy, with adjustments every two weeks, that could shorten the epidemic duration by approximately 7 months. Using International Migrant Stock data and Facebook IP-based mobility data with the Prophet forecasting model, we assessed spillover risk. Four countries were identified as very high risk at the end of July. Compared with the status quo scenario, the optimised control strategy could substantially reduce global importation risk. Enhanced entry screening and preparedness are warranted in neighbouring countries of the DRC in Africa, France in Europe, and Canada in North America.

## Introduction

Ebola virus disease (EVD) is a severe haemorrhagic fever caused by viruses of the genus Ebolavirus, first identified in 1976 near the Ebola River in the Democratic Republic of the Congo (DRC).^1,2^ Classified as a biosafety level 4 (BSL-4) pathogen,^3^ its natural reservoir is primarily believed to reside in wild animals such as bats. Infection can trigger severe haemorrhagic manifestations, with clinical features including fever, vomiting, diarrhoea, and multi-organ failure.^4^ Of the nine Public Health Emergencies of International Concern (PHEIC) declared by the World Health Organization (WHO) to date, three have been Ebola outbreaks—the 2014–2016 West Africa epidemic, the 2018–2020 Kivu epidemic in the DRC, and the 2026 Bundibugyo ebolavirus (BDBV) outbreak in the DRC and neighbouring Uganda.^5^

According to WHO reports, the first confirmed case in the current outbreak was a health-care worker, with symptom onset on 24 April and death on 27 April; however, the precise timing of infection could not be accurately traced, suggesting that the outbreak may have been circulating undetected for several weeks before official confirmation.^6^ Rapid dissemination is thought to have originated from a fatal case on 5 May, yet the outbreak was not laboratory-confirmed as BDBV until 15 May, by which time substantial community transmission had already occurred, posing a sustained and severe threat to global public health.

Estimates of transmission potential for previous African outbreaks, based on classical susceptible–exposed–infectious–recovered (SEIR) compartmental models, have yielded basic reproduction numbers (ℛ_0_) of 2.64 for the 1995 DRC outbreak, 3.54 for the 2000 Uganda outbreak, and 1.53 for the 2007 Uganda outbreak.^7^ The 2014 West Africa epidemic was the largest recorded, with 28 616 cumulative cases and 11 310 deaths across Guinea, Liberia, and Sierra Leone alone.^8^ Modelling estimates for that epidemic yielded ℛ_0_ values of 1.67 (95% CI: 1.59–1.76) for Sierra Leone, 1.82 (95% CI: 1.77–1.83) for Liberia, and 1.49 (95% CI: 1.48–1.50) for Guinea.^9^ The 2018 DRC epidemic resulted in over 2500 infections and more than 1600 deaths.^10^ The 2026 outbreak, which emerged in the DRC and Uganda and whose rapid spread has been traced to a fatal case on 5 May, represents the 17th Ebola epidemic recorded in the country since the virus was first identified in 1976.^11^

A number of mathematical models have been developed to investigate Ebola transmission dynamics and intervention strategies. Legrand and colleagues constructed a six-compartment model encompassing susceptible, exposed, infectious, hospitalised, dead-unburied, and recovered individuals, and were the first to incorporate funeral-related transmission pathways into a quantitative analytical framework.^12^ Feng and colleagues modified this model, arguing that deceased individuals who have been buried should be removed from the population rather than transferred directly to the recovered compartment, thereby rendering the model structure more consistent with the actual transmission process of Ebola.^13^ Wang and colleagues further explored the effects of different stage distributions for the infectious period—including exponential, gamma, and arbitrary distributions—on model behaviour, and used these to evaluate the effectiveness of control strategies such as hospital isolation and timely burial.^14^ Given that Ebola outbreaks frequently occur in regions with scarce medical resources, a model incorporating limited health-care capacity and waning immunity found that increasing the maximum treatment cure rate from 20% to 40% could reduce the peak number of infected individuals by 52.3%; a 10% increase in hospital bed-to-population ratio also reduced the epidemic peak by 29.1%, whereas a reduction in the rate of immunity loss from 60% to 20% decreased the peak infectious burden by 13.2%.^15^

The African continent is undergoing profound and complex population mobility dynamics. According to the 2024 International Migrant Stock estimates from the United Nations Department of Economic and Social Affairs, more than 31 million Africans live outside their country of birth, of whom approximately 21 million reside in another African country.^16^ This high mobility is not only economically driven but also closely intertwined with non-economic factors, including armed conflict, political instability, and climate-induced environmental pressures.^17^ The DRC serves as a critical hub for regional population movement and is also experiencing one of the world’s most severe internal displacement crises, with conflicts and disasters having displaced millions of people.^18,19^ In June 2026, the Africa Centres for Disease Control and Prevention (Africa CDC) and WHO jointly released a response plan for the BDBV outbreak, covering the period from June to November 2026, to strengthen multi-country coordinated responses across surveillance, testing, infection prevention and control, clinical care, community engagement, and cross-border collaboration.^20,21^

To systematically evaluate the transmission dynamics and regional spillover risk of the 2026 BDBV outbreak in the DRC, and to provide quantitative evidence for optimising control strategies in resource-limited settings, we developed a susceptible-exposed-infectious-deceased-recovered (SEIDR) transmission dynamics model that integrates three categories of interventions—public self-protection, treatment and convalescence, and safe burial—within a unified framework. Through sensitivity analyses across multiple candidate start dates, we retrospectively inferred the most likely start date of sustained transmission, calculated the basic reproduction number and its components, and quantified the relative contributions of living-case-mediated and corpse-mediated transmission pathways. We further derived time-varying intervention strategies based on optimal control theory and compared their performance against the status quo and projected scenarios. Concurrently, we used International Migrant Stock data and Facebook IP-based mobility data to assess the risk of spillover to other countries globally. Collectively, these analyses aim to inform the development of dynamic, evidence-based control strategies and cross-border coordination mechanisms for the DRC and the broader international community.

## Methods

### Compartmental modelling framework

Building upon the classical SEIR framework, and incorporating the specific transmission characteristics of Ebola virus and the public health realities in the affected region, we extended the model to include a deceased compartment (D), resulting in a SEIDR structure (supplementary Fig. S1). Traditional burial practices in eastern DRC involve farewell ceremonies, and bodies are typically not cremated. Given that Ebola virus remains highly infectious after death, burial-related contacts constitute a major transmission pathway. Accordingly, the effective force of infection, *λ*, was defined as arising from contact with both living infected individuals and improperly handled deceased bodies, expressed as:

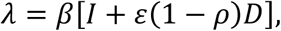

where *β* denotes the effective transmission rate per unit time from a single infected individual to susceptible contacts; ε is a modulating factor representing the relative transmissibility of deceased bodies compared with living infected individuals; and (1 − *ρ*) is the proportion of deceased bodies that are not safely managed. Here, *ρ* denotes the safe burial rate, defined as the proportion of cadavers that are properly buried or disinfected such that they no longer pose a transmission risk.

Three categories of interventions were incorporated to reflect the realistic context of outbreak response: *θ*, the transmission reduction coefficient resulting from public self-protective measures adopted following health education; *ρ*, the safe burial rate as defined above; and *μ*, the recovery rate of infected individuals receiving effective treatment, representing the effect of pharmaceutical interventions on epidemic containment. The structure of the model and the transitions between compartments are described by the following system of transmission dynamics equations (1), with the constraint that *r* + *f* + *μ* ≤ 1.

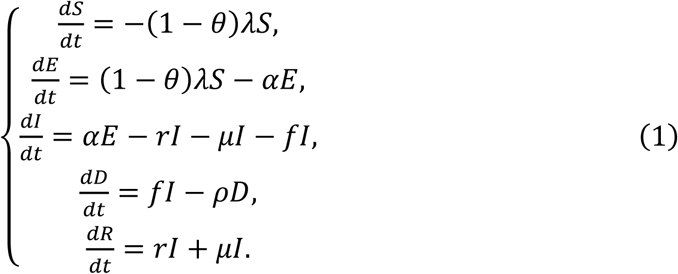

### Data sources and parameter estimation

All data used in this study were obtained from publicly available databases and official reports. Daily cumulative confirmed case counts for the DRC were extracted from WHO’s *Daily epidemiological update on acute public health events: Ebola disease—Bundibugyo virus*,^22^ compiled and provided by the Democratic Republic of the Congo Public Health Emergency Operations Centre (COUSP-DRC). We retrieved the complete time-series data from the start of the outbreak up to the study cut-off date (31 July 2026). Population data were obtained from the *World Population Prospects 2024*, published by the United Nations Department of Economic and Social Affairs, Population Division,^23^ covering the total populations of the DRC and potential risk-importation countries. Cross-border mobility data were derived from the *International Migrant Stock 2024: Destination and Origin*, published by the United Nations Department of Economic and Social Affairs,^24^ with monthly allocation weights determined according to reference,^25^ to estimate the volume and spatial distribution of outflows from the DRC to neighbouring countries and major destination countries. Some epidemiological parameters required by the model were derived from published Ebola virus literature, whereas others were estimated by fitting to the real-time epidemic data collected in this study. Specific values and sources are detailed in supplementary Table S1.

### Optimal control formulation and solution

Pontryagin and Boltyanskii established optimal control theory for systems governed by ordinary differential equations.^26^ Pontryagin’s Maximum Principle (PMP) enables the simultaneous achievement of reducing infection burden and improving model response by optimising time-varying control variables, while accounting for control costs. However, conventional PMP-based indirect methods have inherent limitations in solving multi-objective optimal control problems, including difficulty in selecting initial co-state variable values, narrow convergence regions, and the capacity to obtain only a single locally optimal solution. To overcome these shortcomings, we propose a hybrid solution strategy combining PMP with non-dominated sorting genetic algorithm II (NSGA-II). The core principle is to use a global optimisation algorithm to generate high-quality initial guesses, thereby guiding PMP iterations towards rapid convergence to the globally optimal solution.

Within the framework of model (1), and considering multiple public health interventions comprehensively, the objective function was defined as:

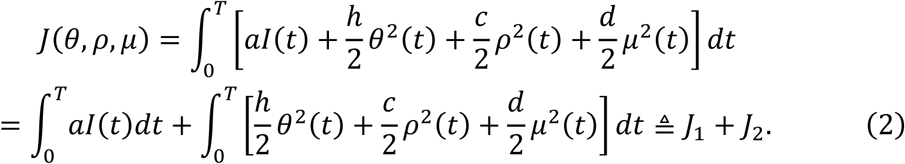

where the constant *a* > 0 is the weight factor for the infectious compartment I, and ℎ, *c*, *d* > 0 are weight factors for the control functions. Our objective was to find a solution *u*^∗^ = (*θ*^∗^, *ρ*^∗^, *μ*^∗^) within the control set

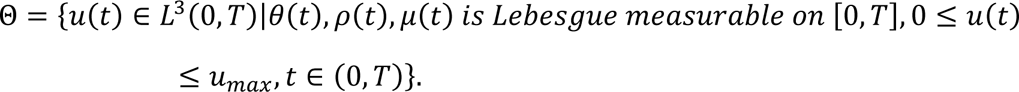

that minimises both the number of infected individuals and the cost of control, i.e.

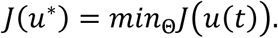

where *T* is the time horizon of the control strategy and *u_max_* is the upper bound of the control functions.

Step 1: Pareto front search

To overcome the inherent limitations of gradient-based indirect methods— namely, sensitivity to initial values and the ability to obtain only a single locally optimal solution—we employed the NSGA-II^27,28^ to generate the Pareto-optimal solution set. This algorithm stratifies population individuals through non-dominated sorting and introduces a crowding distance mechanism to maintain solution diversity, enabling simultaneous search for multiple mutually non-dominated optimal solutions in a single run. After decoding each individual into a piecewise control sequence, the SEIDR state equations (1) were numerically solved using the fourth-order Runge– Kutta method, with the corresponding *J*_1_ and *J*_2_ computed as the fitness vector. The algorithm ultimately outputs a set of non-dominated solutions, i.e. the Pareto front.

Step 2: Ideal point determination

To select a unique optimal solution from the Pareto front, we adopted the ideal point method as a multi-criteria decision-making tool.^29,30^ The normalised Euclidean distance from each solution’s objective vector to the ideal point was calculated as:

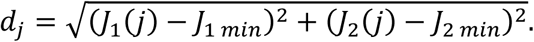

The solution with the minimum distance was deemed the compromise solution achieving the best trade-off between the two conflicting objectives, with its corresponding control strategy being the final compromise-optimal solution (*J*_1 min_, *J*_2 min_).

Step 3: PMP-based refinement

The dynamic constraints described by the state equation *̇x*(*t*) = *g*(*t*, *x*(*t*), *u*(*t*)), *x*(0) = *x*_0_, were incorporated into the optimisation framework together with the control variables. Based on the piecewise constant control strategy corresponding to the compromise-optimal solution, we decoded it to serve as the initial guess for the co-state variable values, thereby transforming the original optimal control problem into a boundary-value problem consisting of state equations, adjoint equations, and optimality conditions, to achieve high-precision numerical solutions for the optimal control trajectories.

The Hamiltonian function was defined as:

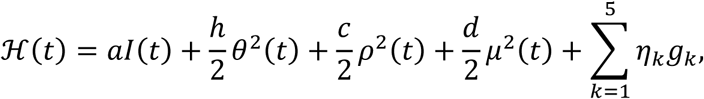

where *g_k_*, (*k* = 1, … ,5) are the right-hand side functions of model (1), and *η_k_*, (*k* = 1, … ,5) denote the adjoint variables corresponding to the state variables of model (1).

Applying the first condition of PMP yields the optimal control functions satisfying:

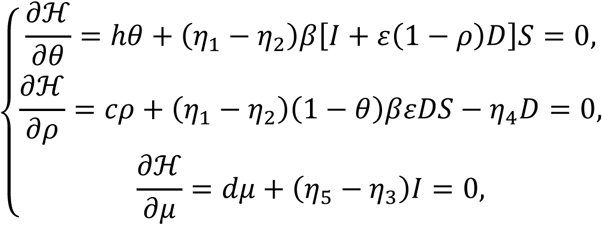

Solving gives:

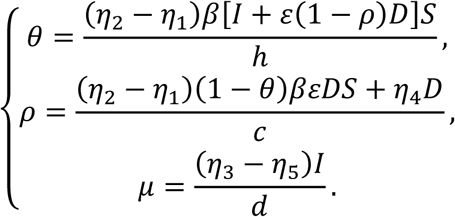

The expressions for the optimal control solutions of this system are:

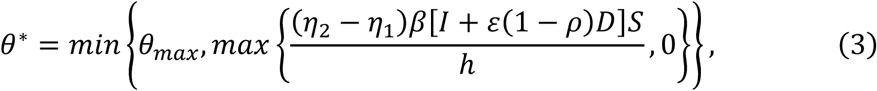

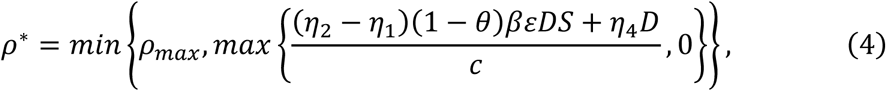

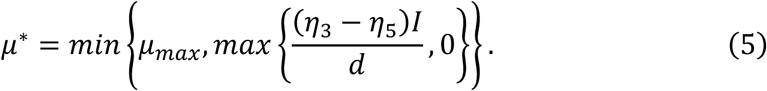

Applying the second condition of PMP yields the adjoint system:

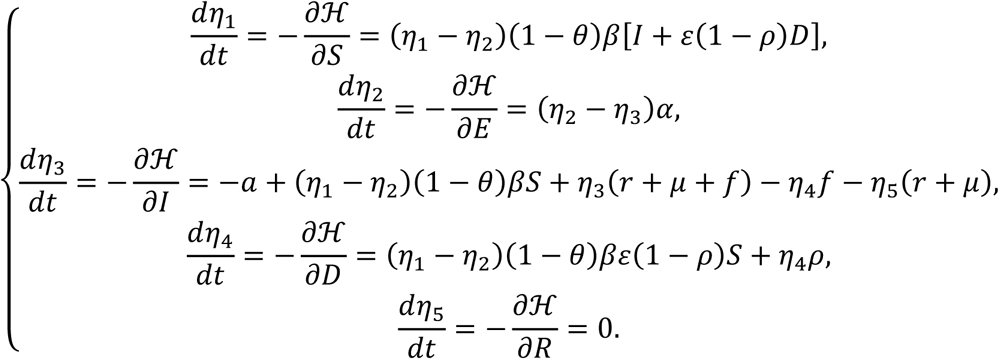

with terminal conditions *η_k_*(*T*) = 0, (*k* = 1, … ,5). A strategy combining gradient descent with Newton iteration was employed, alternately updating the co-state variables and control variables until convergence to a steady state. The final output consists of continuously differentiable optimal control curves and the corresponding epidemic dynamic trajectories.

### Spillover risk estimation with mobility data

To assess the risk of Ebola virus disease spreading beyond the outbreak country (DRC) to other regions, this study incorporated international migration data to evaluate the risk of cross-border spread via population connectivity from the perspective of transnational migration. Assuming homogeneous mixing of the population in the affected country and equal probability of out-migration for all individuals, the number of imported cases from affected country *e* to destination country *b* by month *t* was assumed to follow a binomial distribution:^31,32^

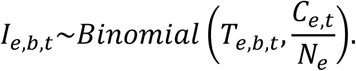

where *T_e_*_,*b*,*t*_ is the cumulative number of out-migrants from affected country *e* to destination country *b* from epidemic onset to month *t*, *C_e_*_,*t*_ is the cumulative number of confirmed cases in affected country *e* as of month *t*, and *N_e_* is the total population of affected country *e*. The use of cumulative cases rather than active infections represents a worst-case scenario assumption, in which all reported infected individuals are considered potentially infectious. Accordingly, the probability of at least one imported case occurring in destination country *b* as of month *t* is:

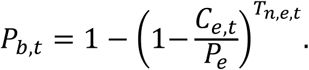

This probability serves as a relative measure of importation risk for each country, with higher values indicating greater risk.

We used the annual International Migrant Stock as a baseline measure of population connectivity strength. To further estimate the monthly proportion of outflows from the DRC to neighbouring and major destination countries in 2026, we referred to reference 25, which provided monthly out-migration data from the DRC based on Facebook IP addresses for the period 2019–2022. We applied the Prophet forecasting model developed by Facebook Meta to capture inherent annual seasonal patterns in migration behaviour and automatically detect trend change points. The mean absolute percentage error (MAPE) was used to evaluate model performance. For countries with MAPE below 50%, the Prophet-generated monthly predictions for 2026 were used as estimates of monthly outflow proportions; for countries with poor predictive performance, the actual monthly migration values from 2022 were used as substitutes to reduce the confounding effects of the COVID-19 pandemic on population mobility.

## Results

### Epidemic tracing and current spillover risk

The first confirmed case of this outbreak was reported on 24 April, with genetic sequencing suggesting that sporadic cases may have emerged as early as February, indicating that the virus had been circulating undetected for several weeks before official confirmation. To investigate the timing of the transition from sporadic cases to sustained transmission, we set the possible start date of sustained transmission to range between 24 March and 24 April, generating 32 candidate start-date scenarios for sustained transmission, each of which was subjected to model fitting. Model fitting was performed using the time-series data of cumulative confirmed cases from 18 May 2026, when WHO officially released information on the outbreak, to 31 July 2026. During the observation period, cumulative confirmed cases increased from 33 to 3,674. Across all 32 candidate start dates for sustained transmission, 28 March was identified as the optimal fitting start date of sustained transmission (Fig. 1a), with an R² of 0.7586, indicating good model fit. Start dates between 31 March and 3 April also yielded good fitting performance, with R² values exceeding 0.7 and ranking among the top across all evaluation metrics (Fig. 1b–e). Goodness-of-fit was evaluated using three metrics: the coefficient of determination (R²), root mean square error (RMSE), and corrected Akaike information criterion (AICc) (supplementary Fig. S3).

**Fig. 1.**
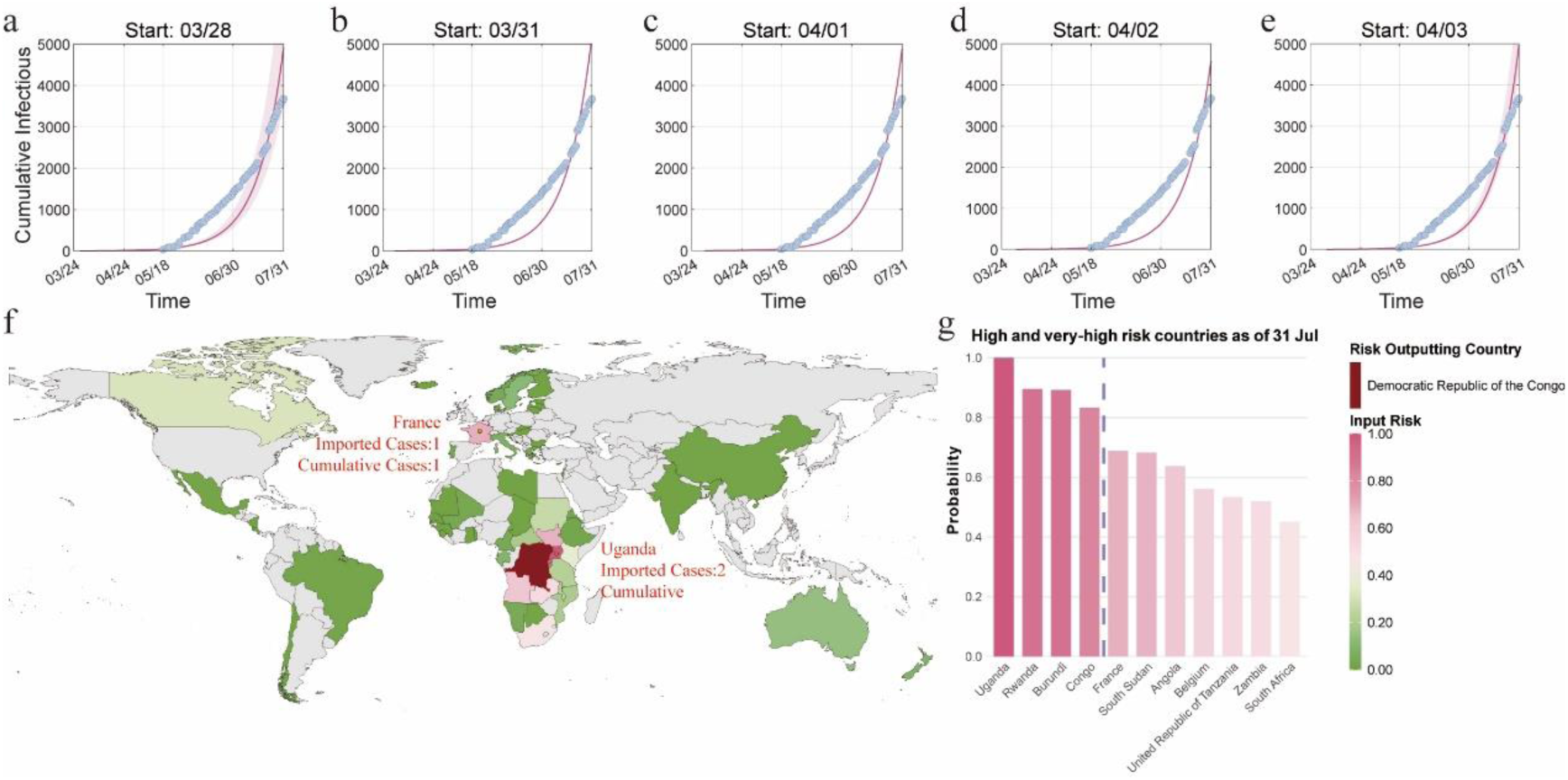
Model fitting of the start date of sustained transmission and cross-border spillover risk as of 31 July. (a–e) Comparison of fitted epidemic curves under different start dates of sustained transmission (R²>0.7) with reported data; (f) geographic distribution of exportation risk from the DRC as of 31 July, estimated from International Migrant Stock data. The outbreak country (DRC) is highlighted in dark red; other countries are shaded from green to pink according to importation risk from highest to lowest, with darker green indicating lower risk. (g) Ranking of very-high-risk and high-risk countries as of 31 July; the purple dashed line indicates the boundary between the two risk tiers.

The outbreak in the DRC remains severe and has already resulted in cross-border transmission. According to reported data, Uganda has reported two imported cases with 20 cumulative confirmed cases domestically; France has reported one imported case with no local secondary transmission. Based on the normal distribution of importation risk probabilities as of end-July, risk was stratified into four levels: 0 < *P_b_*_,*t*_ < 16.38% as low risk, 16.38% ≤ *P_b_*_,*t*_ < 43.68% as moderate risk, 43.68% ≤ *P_b_*_,*t*_ < 70.98% as high risk, and 70.98% ≤ *P_b_*_,*t*_ < 1 as very high risk (supplementary Fig. S4). As of 31 July, a total of 59 countries faced importation risk (*P*(b,t) > 0), including 29 in Africa, 17 in Europe, four in Asia, four in North America, three in South America, and two in Oceania (Fig. 1f). Four countries were classified as very high risk (Fig. 1g): Uganda, Rwanda, Burundi, and the Republic of the Congo. Seven countries were classified as high risk—France, South Sudan, Angola, Belgium, the United Republic of Tanzania, Zambia, and South Africa—indicating substantial importation risk that warrants enhanced entry screening and other control measures.

### Reproduction number and intervention thresholds

The basic reproduction number is a threshold quantity that determines whether a disease can successfully establish transmission and become endemic. It is defined as the average number of secondary infections generated by a single infected individual over the course of their infectious period, in the presence of external interventions. When ℛ_0_ > 1, the disease can become endemic; when ℛ_0_ < 1, the disease will gradually be eliminated. Defining the disease-free equilibrium (DFE) as (*S*_0_, 0,0,0,0) when *I* = 0, and using the next-generation matrix approach, the transmission matrix ℱ and transition matrix V are given by:

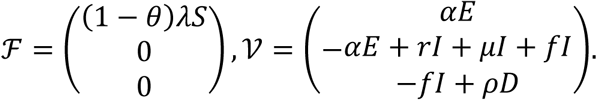

The Jacobian matrices of ℱ and V at the DFE are:

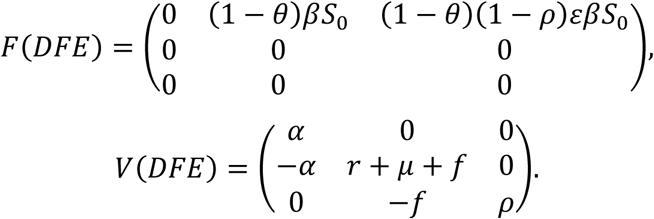

The basic reproduction number is the spectral radius of the matrix *FV*^−1^, yielding:

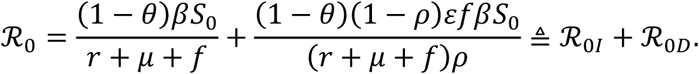

In this decomposition, ℛ_0*I*_ represents the average number of secondary infections generated by an infected individual through effective contacts with a proportion (1 − *θ*) of susceptible individuals at transmission rate *β* over the mean infectious period 1/(*r* + *μ* + *f*). ℛ_0*D*_ represents the average number of secondary infections generated by the body of an infected individual who dies with probability *f*, whose remains are not safely managed (with probability 1 − *ρ*) and thus remain infectious, over the mean time to safe burial 1/*ρ*, through effective contacts with a proportion (1 − *θ*) of susceptible individuals at transmission rate *εβ*.

Based on parameter estimation using cumulative confirmed case data up to 30 June 2026, with parameters estimated via Markov chain Monte Carlo (MCMC) methods, the basic reproduction number for this outbreak was estimated as ℛ_0_ =1.83 (95% CI: 1.81–1.84), of which ℛ_0*I*_ =1.14 (62.18%) and ℛ_0*D*_ =0.69 (37.82%). These findings indicate that the outbreak is in an expanding phase and that, in the absence of interventions, case numbers would continue to rise. It is therefore necessary to further evaluate the impact of the three categories of control measures incorporated in the model on epidemic progression. Given that *ρ* appears in the denominator of the expression for ℛ_0*D*_, where the component is undefined when *ρ* = 0, in subsequent sensitivity analyses, the ranges for *θ* and *μ* were set to [0, 1], and the range for *ρ* was set to [0.1, 1].

When other measures were maintained at baseline levels, higher rates of public self-protection following health education (*θ*) were associated with declining trends in ℛ_0_, ℛ_0*I*_, and ℛ_0*D*_. When *θ* exceeded 58.12%, ℛ_0_ fell below 1. The suppressive effect of *θ* on human-to-human transmission was more pronounced, with ℛ_0*I*_ <1 when *θ* >32.65%, indicating that this measure can effectively interrupt transmission chains driven by infected individuals (Fig. 2a). For pharmaceutical interventions (*μ*), the effect on the deceased-body-mediated transmission pathway was relatively weak, with ℛ_0_ <1 achieved only when *μ* >98.46%. However, *μ* exerted a more substantial effect on interrupting human-to-human transmission, with ℛ_0*I*_ <1 when *μ* >43.31%, suggesting that improving treatment coverage even in resource-limited settings can effectively reduce transmission intensity in the population (Fig. 2b). By contrast, increasing the safe burial rate (*ρ*) had a limited effect on ℛ_0*I*_ and overall ℛ_0_, but substantially reduced secondary transmission resulting from unsafe burials; when *ρ* >20.09%, ℛ_0*D*_ <1, indicating that safe burial practices can effectively eliminate the contribution of the deceased pathway to epidemic amplification (Fig. 2c).

**Fig. 2.**
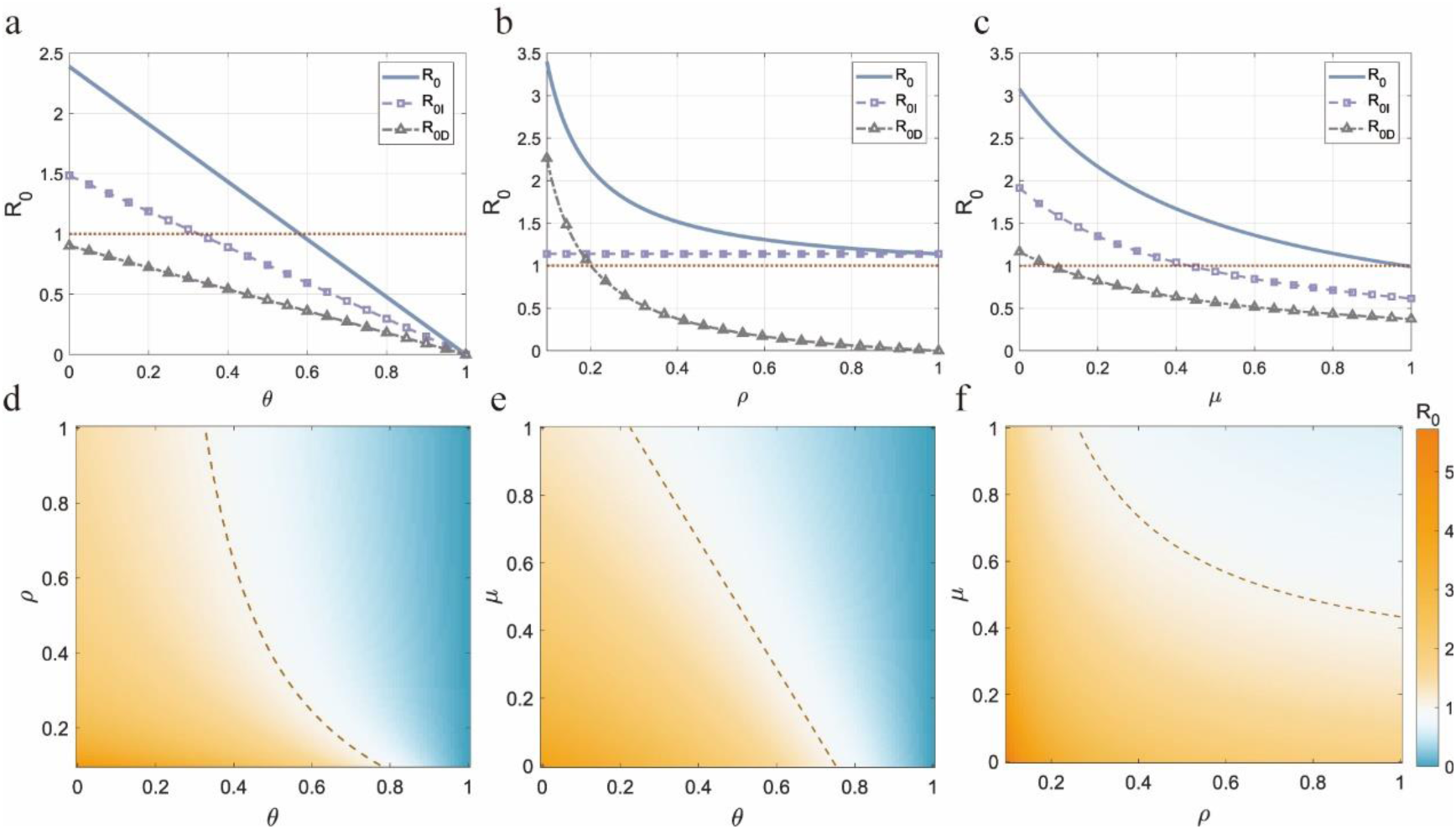
Sensitivity analysis of the basic reproduction number (R_0_) and its components—living-case-mediated transmission (R_0*I*_) and deceased-body-mediated transmission (R_0*D*_)—to the three control measures *θ*, *ρ*, and *μ*. (a–c) One-dimensional curves showing ℛ_0_, ℛ_0*I*_, and ℛ_0*D*_ as functions of public self-protection rate *θ*, safe burial rate *ρ*, and treatment recovery rate *μ*, respectively, with the other two measures fixed at baseline levels; red dashed lines indicate the threshold value of 1. (d–f) Two-dimensional heatmaps showing ℛ_0_, ℛ_0*I*_, and ℛ_0*D*_ as functions of simultaneous variations in the other two measures, with the single measure fixed at baseline level; red dashed lines indicate the threshold value of 1.

With any single measure fixed at baseline level, simultaneous intensification of the other two measures effectively reduced the basic reproduction number, though substantial differences in interaction effects and efficacy boundaries were observed across measures. The concurrent enhancement of self-protection rate (*θ*) and treatment recovery rate (*μ*) yielded the most pronounced suppression of ℛ_0_, whereas the combined regulation of *θ* and safe burial rate (*ρ*) was comparatively weakest (Fig. 2d–f). Notably, when relying on a single intervention alone, the basic reproduction number could reach values above 5 in some parameter regions, further underscoring the necessity and practical importance of multi-measure synergistic interventions in outbreak control.

### Optimal control and projected spillover risk

To achieve effective outbreak control under resource-limited conditions, we pursued the dual objectives of minimising both control costs and infection burden, with weight coefficients set as *a* = 2, ℎ = 1, *c* = 1.5, and *d* = 1. Had time-varying optimal control been applied from 28 March (the model-estimated start date of sustained transmission), the simulated cumulative number of infections as of 31 July would have been approximately 135 cases, representing a 96.33% reduction compared with the 3,674 cases actually reported during the same period (Fig. 3a). Although the explicit solutions of the three optimal control functions are mathematically derived theoretical optima, their simulation results based on actual epidemic data can provide quantitative references for dynamic resource allocation. We suggest that control intensity be reassessed and adjusted approximately every 2 weeks according to the evolving epidemic trajectory—burial measures could be moderately relaxed in subsequent phases, whereas personal protective measures should cover the entire population and treatment resources should be maintained at a relatively high intensity (Table 1 and supplementary Fig. S5). Among them,

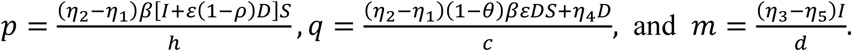

**Fig. 3.**
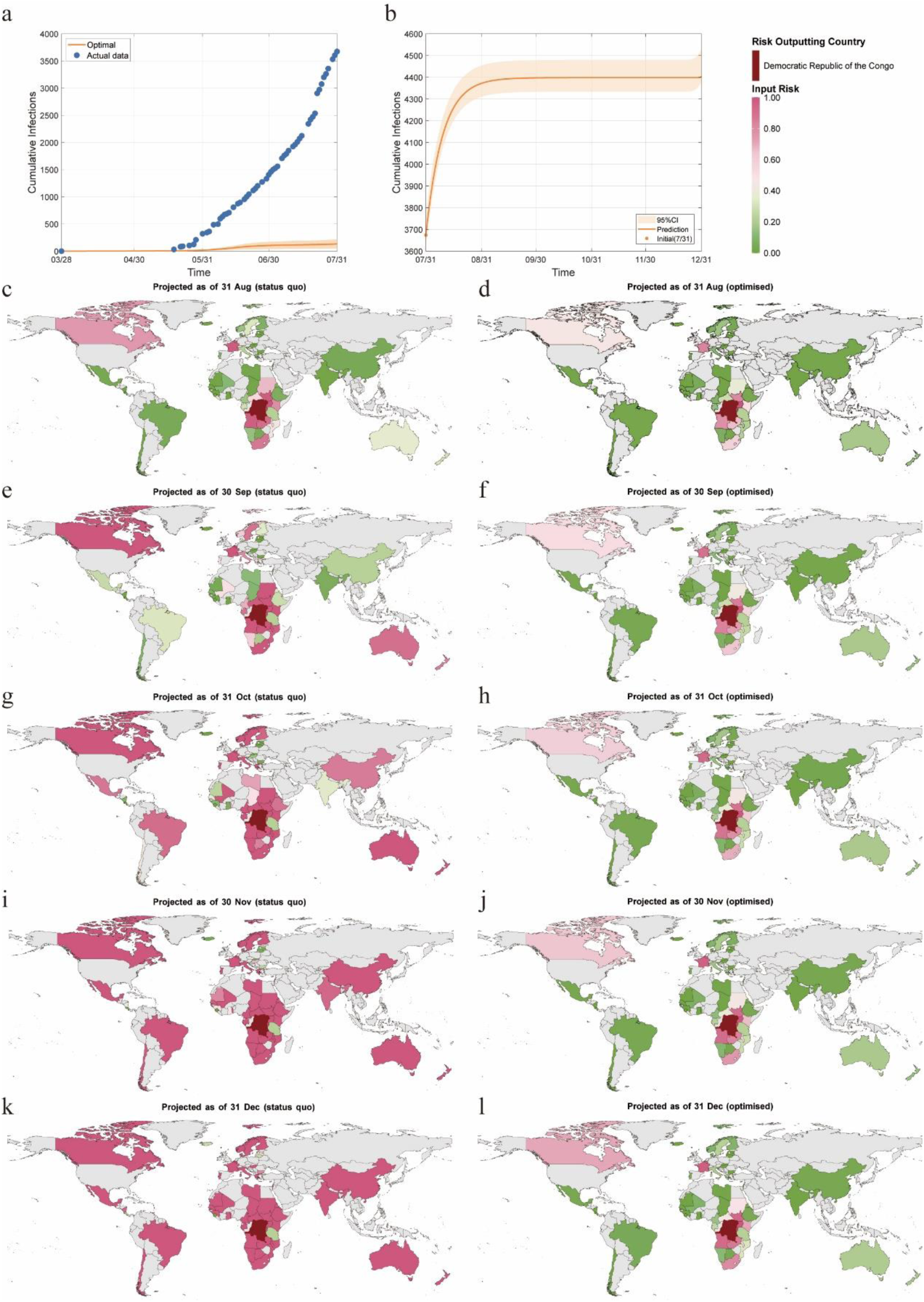
Fitting and projection of cumulative infections under the combined PMP and NSGA-II optimisation , and heatmaps of spillover risk under different control scenarios from August to December 2026. (a) Comparison of model-fitted cumulative infections from epidemic onset to 31 July with reported data. Blue circles denote daily reported cumulative confirmed cases; the red curve shows the simulated cumulative infection trajectory under the optimal control strategy. (b) Projected cumulative infections from 31 July to 31 December under the optimal control strategy, with the actual epidemic size as of 31 July as the initial state. (c–l) Geographic distribution of spillover risk under the status quo control scenario (c, e, g, i, k) and the optimised control scenario (d, f, h, j, l) from August to December 2026.

**Table 1.**
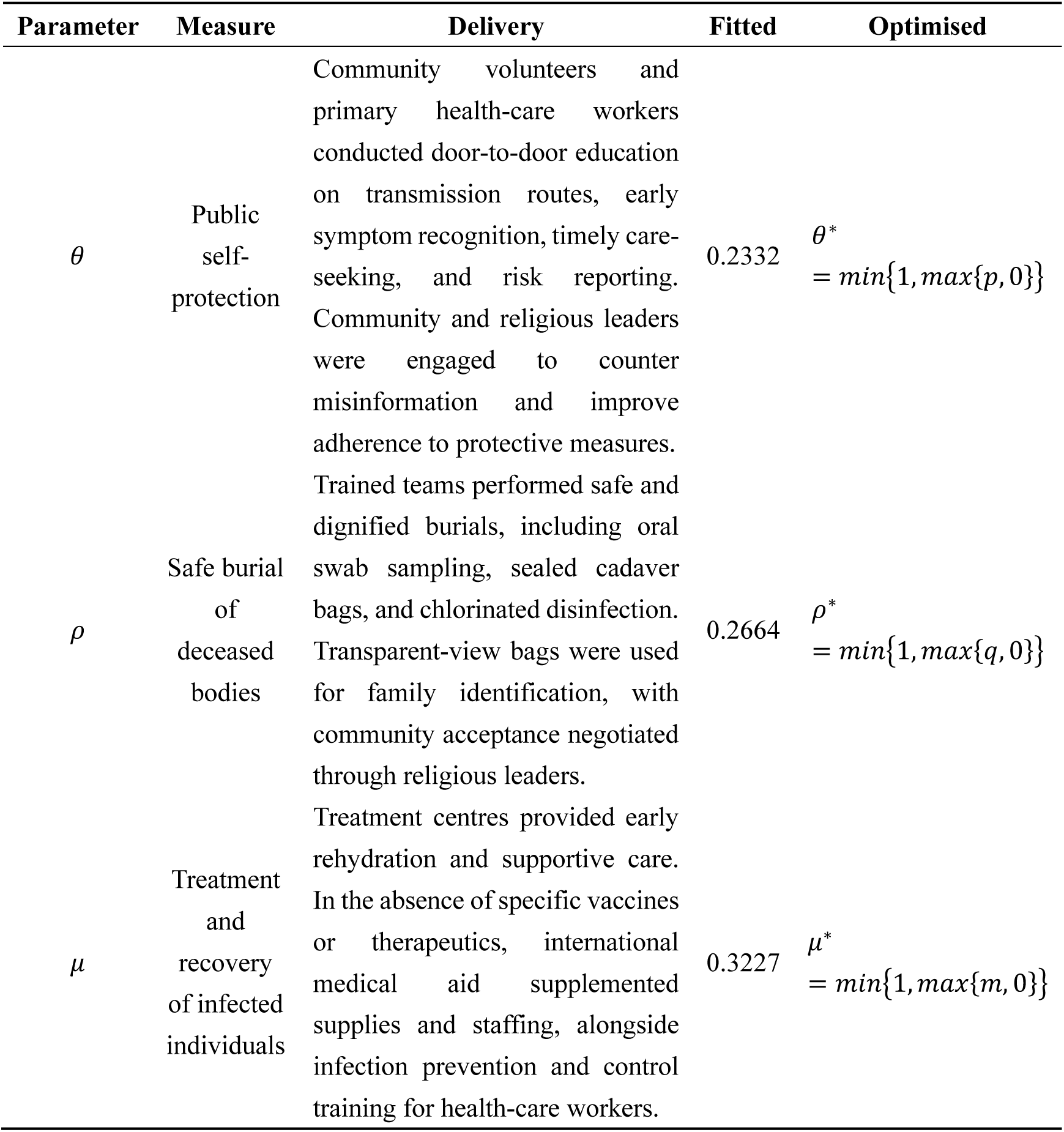
Parameter definitions, implementation approaches, and control intensities for the three intervention categories in the model.

Furthermore, taking the actual epidemic size as of 31 July as the initial state, we projected the epidemic trajectory under the optimal control functions. The projection indicated that cumulative infections would reach 4,372 (95% CI: 4,300–4,450) by 31 August and 4,397 (95% CI: 4,325–4,474) by 30 September, after which the epidemic would enter a plateau phase, with only one additional case by 31 October, suggesting that the outbreak would have been effectively contained by the end of September (Fig. 3b). By contrast, under the status quo control scenario, the epidemic was projected to continue rising until the end of April 2027 (supplementary Fig. S6). The optimal time-varying strategy not only substantially reduced the final epidemic size but also shortened the epidemic duration by approximately 7 months, with significant public health implications.

Comparison of the status quo control scenario with the optimised strategy revealed that optimal control could substantially reduce global importation risk. Under the status quo scenario, the number of very-high-risk countries would increase to 13 by the end of August, with France, South Sudan, Angola, Belgium, the United Republic of Tanzania, Zambia, South Africa, Kenya, and Canada newly added compared with end-July; four countries—Sudan, Malawi, the Central African Republic, and Mozambique—would be classified as high risk (Fig. 3c). By contrast, under the optimised control scenario, eight countries would be at very high risk by the end of August, with France, South Sudan, Angola, and Belgium newly added; five countries would be at high risk, with Kenya and Canada newly added in addition to those not escalating to very high risk (Fig. 3d).

The divergence between the two scenarios widened progressively over time. Under the status quo scenario, the number of very-high-risk countries would increase month by month—to 22 by end-September, 39 by end-October, 47 by end-November, and 53 by end-December. Under the optimised control scenario, by contrast, very-high-risk countries would number ten by end-October and only 11 by end-December (Fig. 3e–l), with high-risk and very-high-risk countries predominantly concentrated in DRC’s neighbouring countries, France, and Canada, substantially mitigating the global public health impact. These findings indicate that, in the coming months, neighbouring countries of the DRC in Africa, France in Europe, and Canada in the Americas face elevated importation risk, necessitating enhanced entry screening and preparedness.

## Discussion

In 2026, the BDBV outbreak was formally confirmed only after a health-care worker in Mongbwalu, Ituri Province, developed severe illness and died; the timing of infection could not be traced, and confirmation relied on laboratory testing following the death of this index case.^33^ Genetic sequencing results suggested that sporadic cases may have emerged as early as February, with some early cases misdiagnosed as malaria or typhoid fever owing to atypical clinical presentations;^34^ however, sustained large-scale transmission had not yet occurred at that time. To investigate the timing of the transition from sporadic cases to sustained community transmission, we assumed that sustained human-to-human transmission began approximately 1 month before the first detected case on 24 April. Our model identified 28 March as the optimal fitting start date of sustained transmission, with the period 31 March to 3 April as the most likely onset window, suggesting that the outbreak had been circulating undetected for approximately 4 weeks before the death of the first confirmed case on 24 April.

BDBV has previously caused two outbreaks. The 2007 outbreak in Uganda reported 116 confirmed cases with 39 deaths, with a basic reproduction number estimated at 1.53;^35^ the 2012 outbreak in the DRC reported 77 cases.^36^ Our model fitting estimated a basic reproduction number of 1.83 (95% CI: 1.81–1.84) for the current outbreak as of 30 June 2026, indicating higher transmission intensity than previous BDBV outbreaks. In terms of outbreak magnitude, as of 11 August 2026, a cumulative total of 4,566 confirmed cases and 2,128 deaths had been reported, surpassing the combined total of all previous BDBV outbreaks. By comparison with the trajectory of the 2014–2016 West Africa Ebola epidemic, the sustained transmission trend in the DRC is concerning.

Like other members of the genus *Ebolavirus*, BDBV is primarily transmitted through direct contact with infected body fluids, contaminated objects, and unsafe burial practices. In terms of case fatality rates, Zaire ebolavirus has the highest fatality rate at 66.6% (95% CI: 55.9–76.8), followed by Sudan ebolavirus at 48.5% (95% CI: 38.6–58.4),^37^ whereas BDBV has a comparatively lower fatality rate. Through decomposition of the basic reproduction number, we quantified the relative contributions of living-case-mediated transmission (62.18%) and deceased-body-mediated transmission (37.28%). At the intervention level, public self-protection following health education was the most effective measure; when 58.12% of susceptible individuals adopted protective behaviours, the transmission chain could be effectively interrupted. Pharmaceutical interventions such as hospitalisation and treatment were also effective in controlling case-mediated transmission; when 43.31% of infected individuals received effective treatment and recovered, the transmission pathway via living cases could be effectively interrupted. Safe burial of deceased bodies also had substantial control value; when 20.09% of cadavers were promptly disinfected and buried without large funeral gatherings, the corpse-mediated transmission pathway could be effectively blocked. Furthermore, the adoption of time-varying optimal control not only reduced cumulative infections by 96.33% by the end of July but also substantially reduced the final epidemic size and shortened the epidemic duration by approximately 7 months.

WHO declared the current outbreak a Public Health Emergency of International Concern on 17 May 2026, assessing the domestic risk in the DRC as “very high” and the regional risk as “high”.^17^ By the end of July 2026, three imported cases had been reported outside the DRC—two in Uganda, with 20 confirmed cases and two deaths reported within Uganda, most linked to cross-border contact—and one in France, involving a health-care worker returning from the DRC who self-reported symptoms and entered isolation upon arrival.^22^ Our migrant-stock-based risk assessment identified four very-high-risk and seven high-risk countries by the end of July, two of which had already reported imported cases, suggesting that Rwanda, Burundi, and the Republic of the Congo also require enhanced surveillance and preparedness.

Furthermore, the implementation of optimised control strategies could effectively reduce spillover risk and prevent broader health consequences. The occurrence of actual imported events validates the effectiveness of our risk assessment model and suggests that the cross-border movement of health-care and humanitarian workers may constitute a specific importation pathway during the early stages of an epidemic.

Currently, no licensed vaccine or specific therapeutic agent is available for BDBV. The existing licensed Ebola vaccine (rVSV-ZEBOV, Ervebo) and therapeutics (Inmazeb, Ebanga) are all designed to target Zaire ebolavirus. For BDBV, WHO launched the first therapeutic trial platform in July 2026 to evaluate MBP-134 (a dual monoclonal antibody) and remdesivir, both as monotherapies and in combination;^38^ the University of Oxford is accelerating the development of the ChAdOx1 BDBV candidate vaccine, with a phase 1 clinical trial initiated in July 2026.^39^ Until specific medical countermeasures become available, WHO emphasises that rapid case identification, isolation, contact tracing, safe burial, and community engagement remain the core components of outbreak control.

Our study has several limitations. The model used the total national population of the DRC as the susceptible population base, without accounting for geographic clustering of the outbreak or the effect of population mobility on contact patterns, which may lead to systematic underestimation of the transmission rate parameter *β*. As of early August 2026, the outbreak in the DRC remains actively transmitting. Although the optimised control functions provide mathematically derived theoretical optima, their operational effectiveness and practical implementation in the actual outbreak context require further validation. Nevertheless, our model results suggest several operationally relevant regulatory rhythms, providing ongoing scientific support for international cross-border collaboration and the formulation of entry screening strategies in high-risk countries, and its analytical framework may serve as a methodological reference for early warning and dynamic response to other Ebolavirus species or emerging haemorrhagic fever outbreaks.

## Supporting information

This file includes: Figures S1 to S9 Tables S1 to S4

## Author contributions

JHL was responsible for investigation, methodology, software, visualisation, writing-original draft. TMC, QPC, JR, and ZYZ contributed to funding acquisition, project administration, and supervision. SJL, YHS, and QPC contributed to formal analysis, visualisation, and validation. All authors contributed to conceptualisation, data curation, and writing-review & editing. All authors had full access to all data in the study and accept responsibility for the decision to submit for publication. TMC, ZYZ, JR, and QPC are the corresponding authors.

## Data Availability

All data used in this study are derived from publicly available sources and have been fully cited in the Data sources section of the manuscript.

