## Supplementary material for "Dynamics, Optimal Control, and Spillover Risk of the 2026 Bundibugyo Ebola Outbreak in the Democratic Republic of the Congo": This file includes: Figures S1 to S9 Tables S1 to S4

Jiahui Li et al.

### This file includes:

Figures S1 to S9

Tables S1 to S4

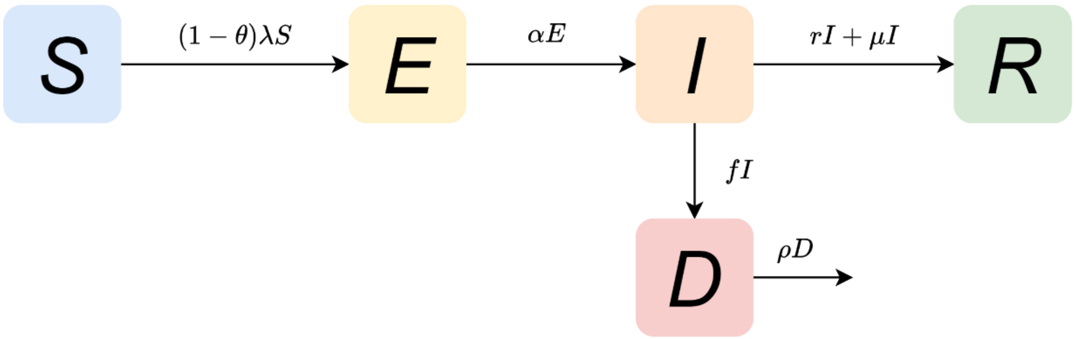


**Fig. S1** Flow diagram of the Ebola virus disease transmission model. S, E, I, D, and R denote susceptible, exposed, infectious, deceased, and recovered individuals, respectively. Susceptible individuals enter the exposed compartment (E) through effective contact with infectious individuals (I) or deceased bodies (D). Exposed individuals are not yet infectious and progress to the infectious compartment (I) after a mean latent period of $1/\alpha$. Among infectious individuals, a proportion r recover naturally and move to the recovered compartment (R); a proportion f die from the disease and enter the deceased compartment (D); and a proportion $\mu$ recover after receiving effective treatment and move to R, with $r + f + \mu \leq 1$. A proportion $\rho$ of deceased bodies are rendered non-infectious through safe burial or disinfection, thereby being removed from the transmission process.

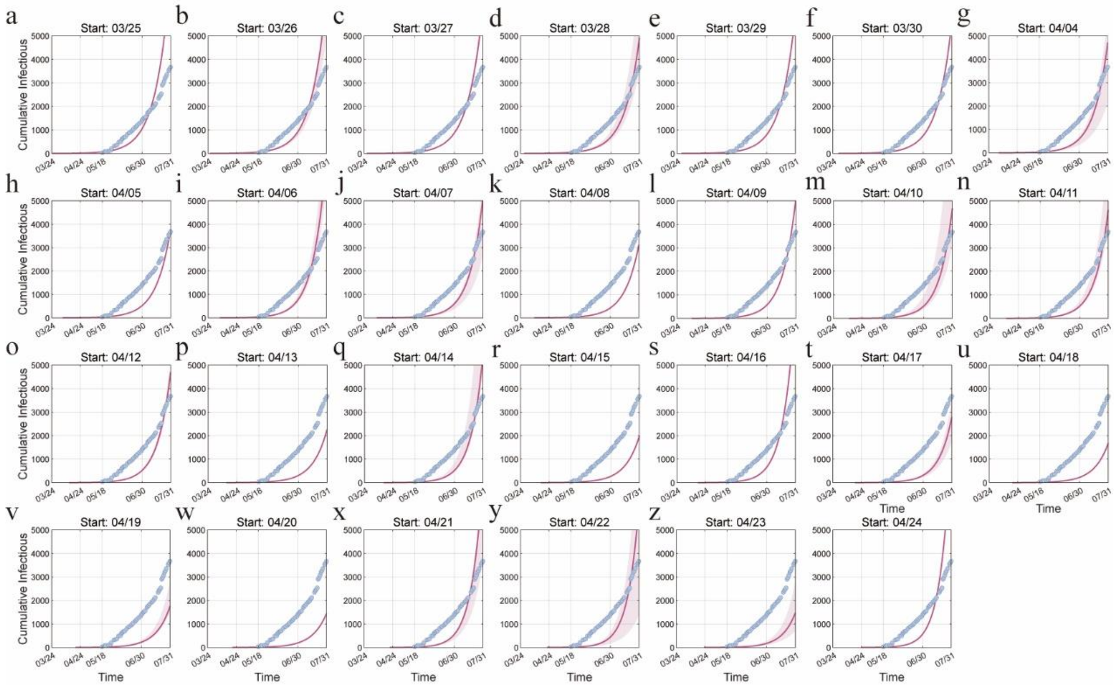


**Fig. S2** Comparison of fitted epidemic curves under different start dates of sustained transmission ($R^2$<0.7) with reported data.

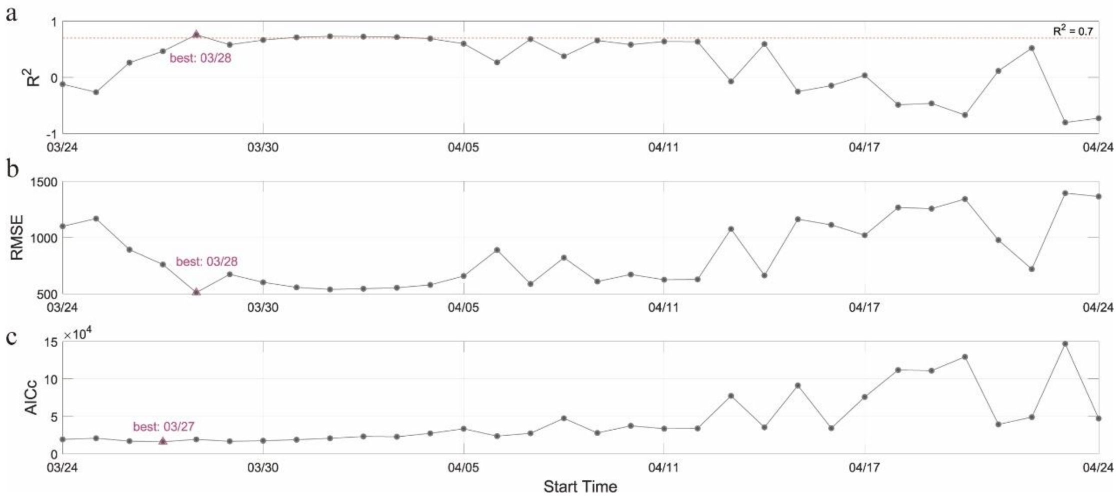


**Fig. S3** Goodness-of-fit metrics for the start date of sustained transmission. (a) coefficient of determination ($R^2$), (b) root mean square error (RMSE), and (c) corrected Akaike information criterion (AICc). Triangles indicate the optimal fitting start date of sustained transmission for each metric.

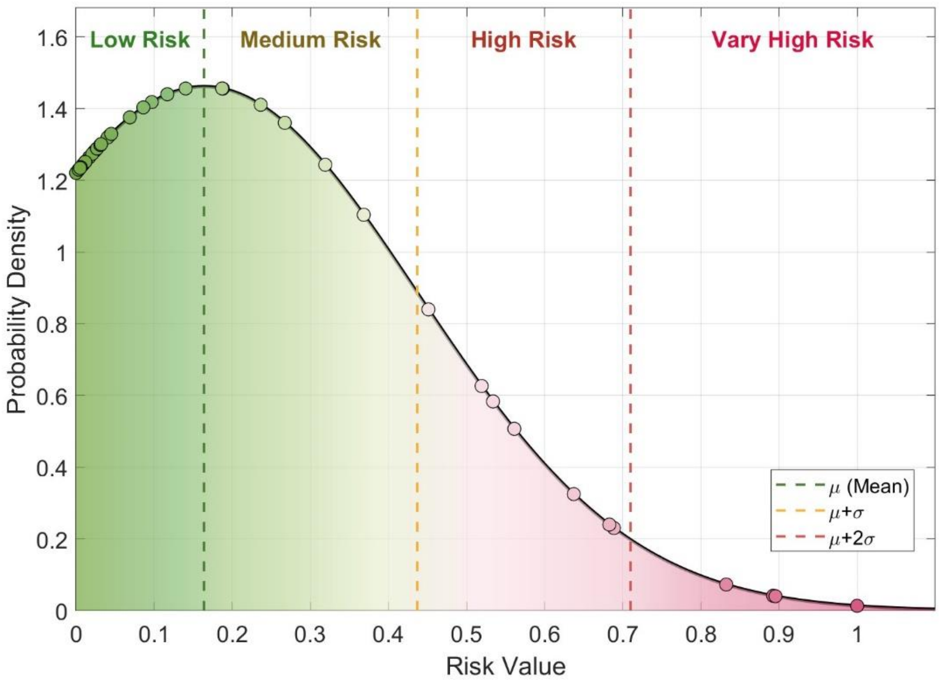


**Fig. S4** Probability density curve of the normal distribution of importation risk probabilities as of end-July. The distribution has a mean ($\mu$) of 16·67% and a standard deviation ($\sigma$) of 28·12%. Based on this distribution, importation risk was stratified into four levels: $0 < P_{b,t} < 16.38\%$ as low risk, $16.38\% \leq P_{b,t} < 43.68\%$ as moderate risk, $43.68\% \leq P_{b,t} < 70.98\%$ as high risk, and $70.98\% \leq P_{b,t} < 1$ as very high risk.

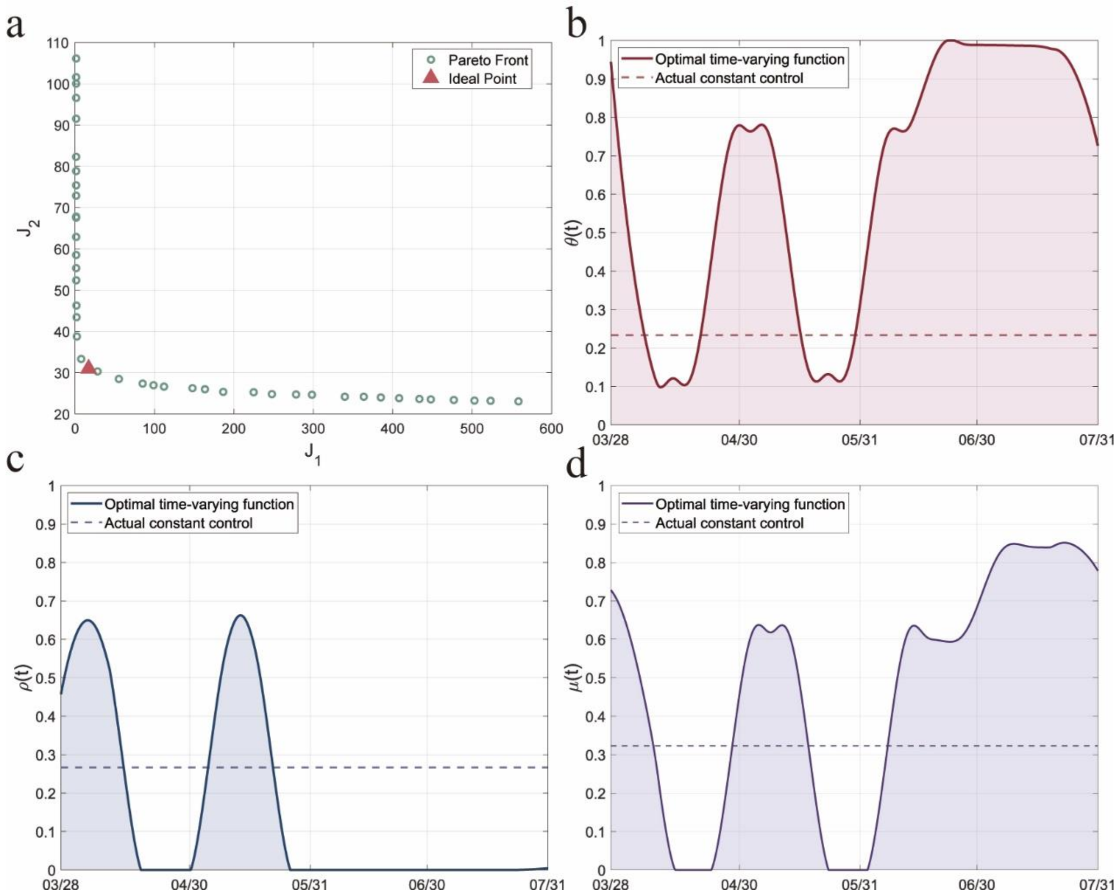


**Fig. S5** Optimal control results from the combined PMP and NSGA-II optimisation. (a) Pareto front and ideal point. Scatter points represent the non-dominated solution set; the red triangle indicates the compromise-optimal solution identified by the ideal point method. (b–d) Comparison of optimal time-varying control strategies with constant control. Solid lines denote the continuous-time optimal control functions derived from PMP; dashed lines indicate the baseline constant control intensity fitted from actual epidemic data.

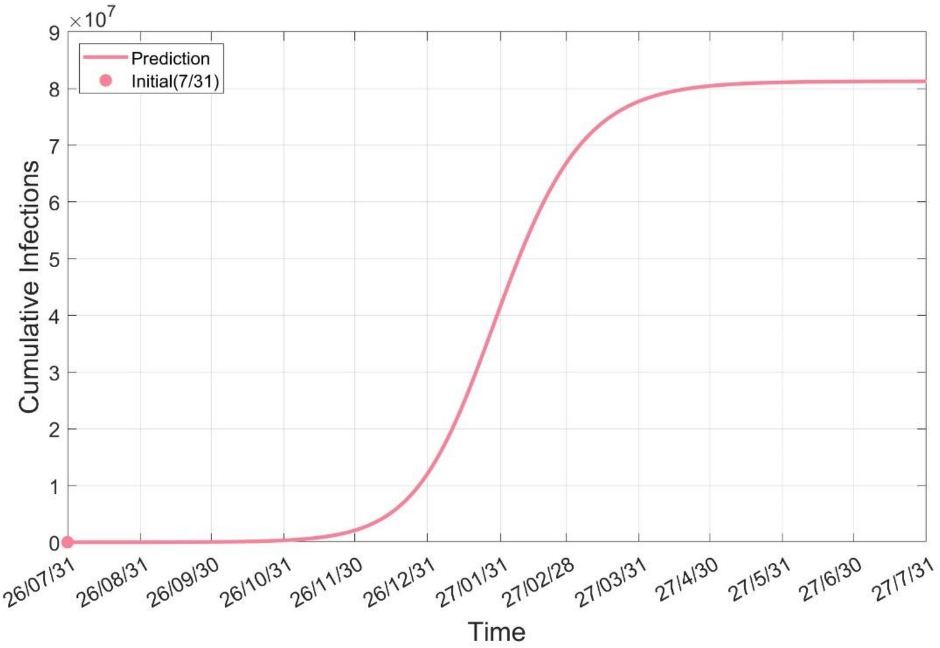


**Fig. S6** Projected cumulative infections from the actual epidemic size as of 31 July 2026 (3 746 confirmed cases) under the status quo control scenario, 31 July 2026 to 31 July 2027.

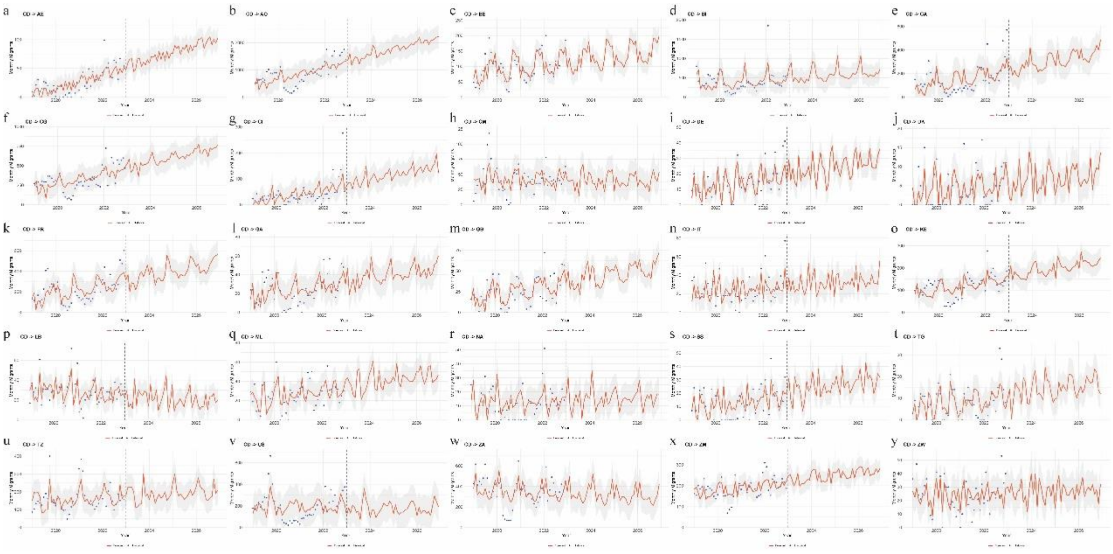

**Fig. S7** Monthly out-migration volumes predicted by the Prophet forecasting model trained on Facebook IP address-based migration data from 2019 to 2022 (for countries with MAPE < 50%).

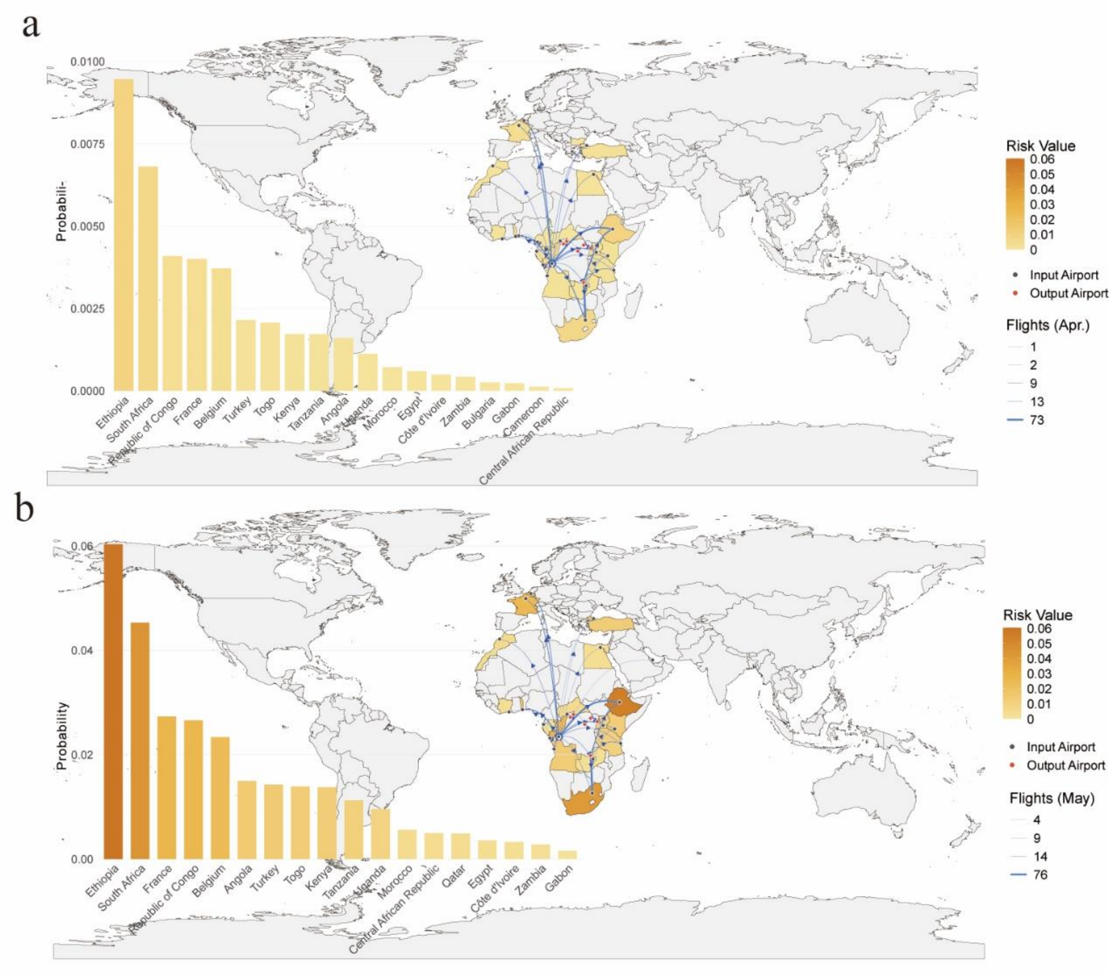


**Fig. S8** Geographic distribution and ranking of all at-risk countries for exportation risk from the DRC based on flight data, April and May 2026. (a) Spatial distribution of the probability of at least one imported case from the DRC via air travel to destination countries in April. Colour intensity reflects risk magnitude, with darker shades indicating higher probabilities. Red dots indicate the seven major international airports within the DRC (Bunia, Gbadolite, Gemena, Isiro, Kinshasa N'Djili International Airport, Kisangani, and Lubumbashi); blue dots indicate airport locations in destination countries; blue directed line segments indicate flight directions, with line thickness reflecting monthly flight frequency. The bar chart on the left ranks countries by importation risk from highest to lowest. (b) Same as (a), for May.

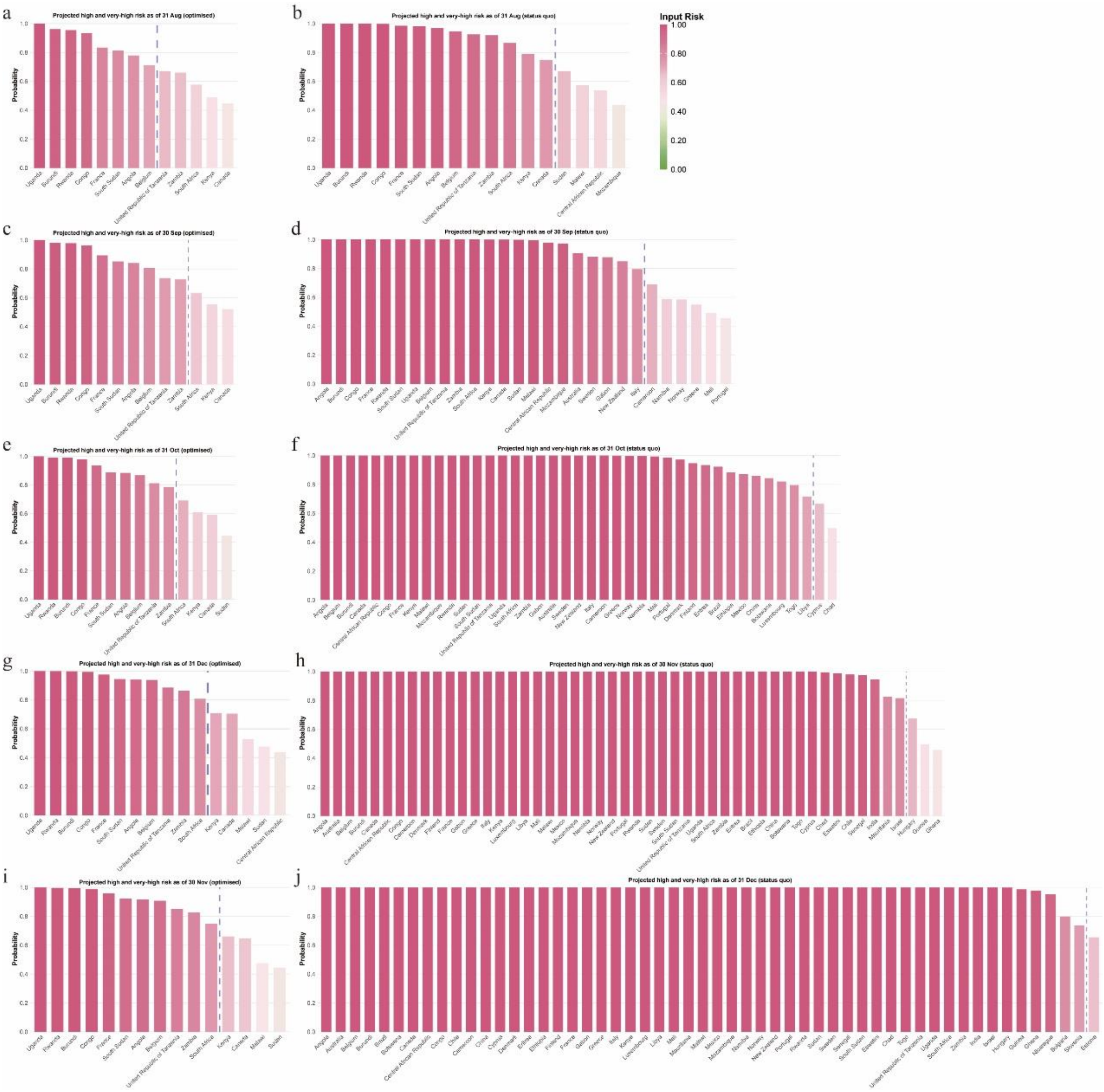


**Fig. S9** Very-high-risk and high-risk countries for spillover risk from the DRC estimated from International Migrant Stock data, August–December 2026. (a, c, e, g, i) Ranking of very-high-risk and high-risk countries under the projected optimised control scenario, as of end-August to end-December, respectively. (b, d, f, h, j) Ranking of very-high-risk and high-risk countries under the projected status quo control scenario, as of end-August to end-December, respectively.

**Table S1** Definition and value of parameters in the transmission dynamics model.

| Parameters | Definition | Value | Unit | Range | Source |
|---|---|---|---|---|---|
| $\beta$ | Effective transmission rate of infected individuals | 1.0823e-08 | $day^{-1}$ | [2.51e-9,1.1e-8] | Fitted |
| $\varepsilon$ | Modulation factor of improperly handled deceased bodies on effective transmission | 0.6692 | NA | [0,1] | Ref. (1, 2) |
| $\alpha$ | Rate of movement out of the exposed compartment | 0.1074 | $day^{-1}$ | [0.1,0.125] | Ref. (1, 3) |

| | | | | | |
|---|---|---|---|---|---|
| $r$ | Natural recovery rate of infected individuals | 0.1438 | $day^{-1}$ | [0.1-0.202] | Ref. (4-6) |
| $f$ | Case fatality rate of infected individuals | 0.33 | $day^{-1}$ | [0.21,0.51] | Ref. (7) |
| $\rho$ | Safe burial rate of deceased individuals | 0.2664 | $day^{-1}$ | [0,1] | Fitted |
| $\theta$ | Rate of self-protection following health education | 0.2332 | $day^{-1}$ | [0,1] | Fitted |
| $\mu$ | Treatment and convalescence rate | 0.3227 | $day^{-1}$ | [0,1] | Fitted |

**Table S2** Importation risk probabilities estimated from International Migrant Stock data, as of 31 July 2026.

| Country | Input risk |
|---|---|
| Angola | 63.71% |
| Australia | 14.03% |
| Belgium | 56.10% |
| Bulgaria | 0.02% |
| Burundi | 89.24% |
| Bolivia (Plurinational State of) | 0.01% |
| Brazil | 1.65% |
| Botswana | 1.06% |
| Canada | 31.90% |
| Central African Republic | 18.78% |
| Congo | 83.23% |
| Chile | 0.46% |
| Cameroon | 4.37% |
| China | 1.22% |
| Costa Rica | 0.00% |
| Cyprus | 0.88% |
| Denmark | 2.30% |
| Estonia | 0.01% |
| Eritrea | 2.51% |
| Ethiopia | 2.07% |
| Finland | 0.40% |
| France | 68.86% |
| Gabon | 9.69% |
| Ghana | 0.02% |
| Guinea | 0.07% |
| Greece | 2.68% |
| Hungary | 0.00% |
| Israel | 0.04% |
| India | 0.31% |
| Iceland | 0.00% |

| | |
|---|---|
| Italy | 6.88% |
| Kenya | 36.83% |
| Liechtenstein | 0.00% |
| Lithuania | 0.01% |
| Luxembourg | 0.00% |
| Latvia | 0.01% |
| Libya | 0.33% |
| Mali | 3.11% |
| Mauritania | 0.23% |
| Malawi | 23.64% |
| Mexico | 1.16% |
| Mozambique | 18.70% |
| Namibia | 4.02% |
| Nicaragua | 0.05% |
| Norway | 4.50% |
| New Zealand | 8.60% |
| Portugal | 3.20% |
| Rwanda | 89.54% |
| Sudan | 26.72% |
| Sweden | 11.68% |
| Slovenia | 0.02% |
| Slovakia | 0.01% |
| Sierra Leone | 0.00% |
| Senegal | 0.33% |
| South Sudan | 68.27% |
| Eswatini | 0.59% |
| Chad | 0.59% |
| Togo | 0.51% |
| United Republic of Tanzania | 53.38% |
| Uganda | 99.99% |
| South Africa | 45.11% |
| Zambia | 51.92% |

**Table S3** Projected spillover risk probabilities under the status quo control scenario, as of end-August to end-December 2026.

| Country | August | September | October | November | December |
|---|---|---|---|---|---|
| Angola | 97.07% | 100.00% | 100.00% | 100.00% | 100.00% |
| Australia | 36.19% | 90.65% | 100.00% | 100.00% | 100.00% |
| Belgium | 94.48% | 100.00% | 100.00% | 100.00% | 100.00% |
| Bulgaria | 0.07% | 0.62% | 4.01% | 24.34% | 79.94% |
| Burundi | 99.95% | 100.00% | 100.00% | 100.00% | 100.00% |
| Bolivia (Plurinational State of) | 0.02% | 0.12% | 0.84% | 5.16% | 26.30% |
| Brazil | 4.52% | 32.45% | 92.27% | 100.00% | 100.00% |
| Botswana | 4.27% | 19.77% | 84.36% | 100.00% | 100.00% |

| | | | | | |
|---|---|---|---|---|---|
| Canada | 74.85% | 99.98% | 100.00% | 100.00% | 100.00% |
| Central African Republic | 53.67% | 97.95% | 100.00% | 100.00% | 100.00% |
| Congo | 99.82% | 100.00% | 100.00% | 100.00% | 100.00% |
| Chile | 1.64% | 8.02% | 42.04% | 97.96% | 100.00% |
| Cameroon | 15.69% | 69.03% | 99.99% | 100.00% | 100.00% |
| China | 4.15% | 22.66% | 85.86% | 100.00% | 100.00% |
| Costa Rica | 0.01% | 0.08% | 0.53% | 3.27% | 17.44% |
| Cyprus | 2.64% | 13.94% | 66.60% | 99.94% | 100.00% |
| Denmark | 7.00% | 36.11% | 97.23% | 100.00% | 100.00% |
| Estonia | 0.03% | 0.17% | 1.95% | 16.89% | 65.53% |
| Eritrea | 6.81% | 34.01% | 93.37% | 100.00% | 100.00% |
| Ethiopia | 6.30% | 28.02% | 88.31% | 100.00% | 100.00% |
| Finland | 7.51% | 34.45% | 94.72% | 100.00% | 100.00% |
| France | 98.48% | 100.00% | 100.00% | 100.00% | 100.00% |
| Gabon | 29.33% | 87.80% | 100.00% | 100.00% | 100.00% |
| Ghana | 0.07% | 0.68% | 6.23% | 45.67% | 97.84% |
| Guinea | 0.25% | 1.27% | 10.22% | 49.54% | 98.72% |
| Greece | 7.26% | 54.88% | 99.93% | 100.00% | 100.00% |
| Hungary | 0.00% | 2.70% | 16.35% | 67.35% | 99.84% |
| Israel | 0.81% | 4.04% | 23.61% | 81.52% | 100.00% |
| India | 0.99% | 5.57% | 34.64% | 94.52% | 100.00% |
| Iceland | 0.01% | 0.06% | 0.58% | 3.56% | 26.68% |
| Italy | 23.22% | 79.57% | 100.00% | 100.00% | 100.00% |
| Kenya | 79.15% | 99.99% | 100.00% | 100.00% | 100.00% |
| Liechtenstein | 0.02% | 0.09% | 0.71% | 5.01% | 28.31% |
| Lithuania | 0.02% | 0.09% | 0.57% | 5.00% | 26.95% |
| Luxembourg | 2.19% | 10.56% | 82.03% | 100.00% | 100.00% |
| Latvia | 0.02% | 0.09% | 0.85% | 6.42% | 31.76% |
| Libya | 1.84% | 12.33% | 71.57% | 99.96% | 100.00% |
| Mali | 10.84% | 49.11% | 99.37% | 100.00% | 100.00% |
| Mauritania | 0.72% | 3.58% | 24.32% | 82.57% | 100.00% |
| Malawi | 57.22% | 99.46% | 100.00% | 100.00% | 100.00% |
| Mexico | 3.41% | 27.03% | 87.21% | 100.00% | 100.00% |
| Mozambique | 43.68% | 97.26% | 100.00% | 100.00% | 100.00% |
| Namibia | 14.47% | 58.97% | 99.89% | 100.00% | 100.00% |
| Nicaragua | 0.15% | 0.94% | 8.10% | 41.13% | 95.27% |
| Norway | 15.49% | 58.48% | 99.90% | 100.00% | 100.00% |
| New Zealand | 31.23% | 84.90% | 100.00% | 100.00% | 100.00% |
| Portugal | 10.00% | 45.56% | 98.52% | 100.00% | 100.00% |
| Rwanda | 99.93% | 100.00% | 100.00% | 100.00% | 100.00% |
| Sudan | 67.13% | 99.84% | 100.00% | 100.00% | 100.00% |
| Sweden | 32.80% | 88.24% | 100.00% | 100.00% | 100.00% |
| Slovenia | 0.06% | 0.41% | 3.62% | 20.64% | 73.59% |
| Slovakia | 0.02% | 0.11% | 0.73% | 4.46% | 37.69% |

| | | | | | |
|---|---|---|---|---|---|
| Sierra Leone | 0.00% | 0.00% | 0.00% | 0.00% | 0.00% |
| Senegal | 1.12% | 6.27% | 39.80% | 97.60% | 100.00% |
| South Sudan | 98.04% | 100.00% | 100.00% | 100.00% | 100.00% |
| Eswatini | 1.64% | 8.00% | 41.97% | 98.76% | 100.00% |
| Chad | 1.63% | 9.98% | 49.65% | 99.46% | 100.00% |
| Togo | 2.08% | 16.51% | 79.61% | 100.00% | 100.00% |
| United Republic of Tanzania | 92.51% | 100.00% | 100.00% | 100.00% | 100.00% |
| Uganda | 100.00% | 100.00% | 100.00% | 100.00% | 100.00% |
| South Africa | 86.56% | 100.00% | 100.00% | 100.00% | 100.00% |
| Zambia | 91.95% | 100.00% | 100.00% | 100.00% | 100.00% |

**Table S4** Projected spillover risk probabilities under the optimised control scenario, as of end-August to end-December 2026.

| Country | August | September | October | November | December |
|---|---|---|---|---|---|
| Angola | 78.01% | 84.07% | 88.45% | 91.69% | 94.05% |
| Australia | 17.53% | 18.33% | 18.33% | 18.33% | 19.73% |
| Belgium | 71.15% | 80.58% | 86.82% | 90.79% | 93.86% |
| Bulgaria | 0.03% | 0.05% | 0.05% | 0.06% | 0.06% |
| Burundi | 96.23% | 98.01% | 99.04% | 99.52% | 99.79% |
| Bolivia (Plurinational State of) | 0.01% | 0.01% | 0.01% | 0.01% | 0.01% |
| Brazil | 1.97% | 3.30% | 3.30% | 3.30% | 4.26% |
| Botswana | 1.85% | 1.86% | 2.40% | 2.64% | 3.23% |
| Canada | 44.69% | 52.17% | 59.24% | 64.76% | 70.49% |
| Central African Republic | 28.12% | 28.25% | 36.84% | 38.69% | 43.74% |
| Congo | 93.30% | 96.24% | 97.90% | 98.82% | 99.36% |
| Chile | 0.71% | 0.71% | 0.71% | 0.81% | 0.81% |
| Cameroon | 7.06% | 9.53% | 11.33% | 12.90% | 15.03% |
| China | 1.80% | 2.17% | 2.53% | 2.88% | 3.26% |
| Costa Rica | 0.01% | 0.01% | 0.01% | 0.01% | 0.01% |
| Cyprus | 1.14% | 1.27% | 1.43% | 1.54% | 1.78% |
| Denmark | 3.07% | 3.75% | 4.59% | 5.33% | 6.48% |
| Estonia | 0.01% | 0.01% | 0.03% | 0.04% | 0.04% |
| Eritrea | 2.98% | 3.49% | 3.49% | 3.49% | 3.73% |
| Ethiopia | 2.75% | 2.77% | 2.77% | 3.17% | 3.76% |
| Finland | 3.29% | 3.54% | 3.78% | 5.84% | 8.96% |
| France | 83.40% | 89.43% | 93.40% | 95.94% | 97.52% |
| Gabon | 13.84% | 16.45% | 19.44% | 22.43% | 25.68% |
| Ghana | 0.03% | 0.06% | 0.08% | 0.13% | 0.14% |
| Guinea | 0.11% | 0.11% | 0.14% | 0.14% | 0.16% |
| Greece | 3.18% | 6.57% | 9.03% | 15.76% | 17.97% |
| Hungary | 0.00% | 0.23% | 0.23% | 0.23% | 0.23% |
| Israel | 0.35% | 0.35% | 0.35% | 0.35% | 0.40% |
| India | 0.42% | 0.49% | 0.56% | 0.60% | 0.68% |
| Iceland | 0.00% | 0.00% | 0.01% | 0.01% | 0.01% |

| | | | | | |
|---|---|---|---|---|---|
| Italy | 10.72% | 12.69% | 14.56% | 16.34% | 19.50% |
| Kenya | 48.96% | 55.36% | 60.87% | 66.07% | 70.79% |
| Liechtenstein | 0.01% | 0.01% | 0.01% | 0.01% | 0.01% |
| Lithuania | 0.01% | 0.01% | 0.01% | 0.01% | 0.01% |
| Luxembourg | 0.94% | 0.95% | 2.22% | 2.62% | 3.35% |
| Latvia | 0.01% | 0.01% | 0.01% | 0.01% | 0.01% |
| Libya | 0.80% | 1.12% | 1.63% | 1.63% | 1.63% |
| Mali | 4.81% | 5.61% | 6.42% | 7.24% | 8.15% |
| Mauritania | 0.31% | 0.31% | 0.36% | 0.36% | 0.36% |
| Malawi | 30.53% | 36.02% | 42.29% | 47.48% | 52.70% |
| Mexico | 1.48% | 2.66% | 2.66% | 2.66% | 3.24% |
| Mozambique | 21.83% | 26.46% | 27.99% | 27.99% | 32.36% |
| Namibia | 6.49% | 7.33% | 8.57% | 10.23% | 11.77% |
| Nicaragua | 0.06% | 0.08% | 0.11% | 0.11% | 0.11% |
| Norway | 6.96% | 7.24% | 8.62% | 8.85% | 8.85% |
| New Zealand | 14.84% | 14.91% | 14.92% | 14.92% | 18.51% |
| Portugal | 4.42% | 5.06% | 5.37% | 6.09% | 6.09% |
| Rwanda | 95.53% | 97.83% | 99.12% | 99.70% | 99.90% |
| Sudan | 37.96% | 42.43% | 44.47% | 44.47% | 47.71% |
| Sweden | 15.68% | 16.72% | 17.04% | 17.67% | 22.83% |
| Slovenia | 0.03% | 0.04% | 0.05% | 0.05% | 0.05% |
| Slovakia | 0.01% | 0.01% | 0.01% | 0.01% | 0.02% |
| Sierra Leone | 0.00% | 0.00% | 0.00% | 0.00% | 0.00% |
| Senegal | 0.48% | 0.55% | 0.66% | 0.78% | 0.87% |
| South Sudan | 81.48% | 85.20% | 88.69% | 92.23% | 94.49% |
| Eswatini | 0.71% | 0.71% | 0.71% | 0.91% | 0.91% |
| Chad | 0.70% | 0.89% | 0.89% | 1.08% | 1.16% |
| Togo | 0.90% | 1.53% | 2.06% | 2.32% | 2.47% |
| United Republic of Tanzania | 67.11% | 73.64% | 81.21% | 85.20% | 88.58% |
| Uganda | 100.00% | 100.00% | 100.00% | 100.00% | 100.00% |
| South Africa | 57.73% | 63.40% | 69.18% | 74.80% | 80.74% |
| Zambia | 66.08% | 72.72% | 78.41% | 82.67% | 86.28% |


1. Luo D, Zheng R, Wang D, et al. Effect of sexual transmission on the West Africa Ebola outbreak in 2014: a mathematical modelling study. Sci Rep. 2019; 9(1): 1653. https://doi.org/10.1038/s41598-018-38397-3.
2. Zhu Q, Asim M, Ullah S, et al. Nonlinear dynamical modeling and control of Ebola involving transmission from hospitalized and deceased populations: a data-driven approach from Sierra Leone. J Appl Math Comput. 2026; 72: 35.
3. Breman JG, Johnson KM. Ebola then and now. N Engl J Med. 2014; 371: 1663-1666.
4. Bogoch II, Creatore MI, Cetron MS, et al. Assessment of the potential for international dissemination of Ebola virus via commercial air travel during the

2014 West African outbreak. Lancet. 2015; 385: 29-35.

5. Djiomba SD, Nyabadza F. Modelling the potential role of media campaigns in Ebola transmission dynamics. J Differ Equ. 2017; 2017: 1-13.
6. Ahmad MD, Usman M, Khan A, Imran M. Optimal control analysis of Ebola disease with control strategies of quarantine and vaccination. Infect Dis Poverty. 2016; 5(1): 72.
7. World Health Organization. Disease Outbreak News; Bundibugyo Virus Disease, Democratic Republic of the Congo and Uganda. https://cdn.who.int/media/docs/default-source/_sage-2026/20260526_public-rra_bvd_democratic-republic-of-the-congo_v2---finalaa5ebe4a-1d41-401d-96aa-89cd766743cb.pdf?sfvrsn=d14d41a3_4&download=true (accessed June 8, 2026).
